# Deaths from cardiovascular disease involving anticoagulants: a systematic synthesis of coroners’ case reports

**DOI:** 10.1101/2021.04.28.21256272

**Authors:** Ali Anis, Carl Heneghan, Jeffrey K. Aronson, Nicholas J. DeVito, Georgia C. Richards

## Abstract

**Background:** The global burden of cardiovascular disease (CVD) is forecast to increase, and anticoagulants will remain important medicines for its management. Coroners’ Prevention of Future Death reports (PFDs) provide valuable insights that may enable safer and more effective use of these agents.

**Aim:** To identify CVD-related PFDs involving anticoagulants.

**Design and Setting:** Retrospective observational study of coronial case reports in England and Wales between 2013 and 2019.

**Method:** We screened 3037 PFDs for eligibility and included PFDs where CVD and an anticoagulant caused or contributed to the death. We descriptively analysed included cases and used content analysis to assess concerns raised by coroners and who responded to them.

**Results:** We identified 113 cardiovascular disease-related PFDs involving anticoagulants. Warfarin (36%), enoxaparin (11%), and rivaroxaban (11%) were the most common anticoagulants reported. Concerns most frequently raised by coroners included poor systems (31%), poor communication (25%), and failures to keep accurate medical records (25%). These concerns were most often directed to NHS trusts (29%), hospitals (10%), and general practices (8%). Nearly two-thirds (60%) of PFDs had not received responses from such organisations, which are mandatory under regulation 28 of the Coroners’ (Investigations) Regulations 2013. We created a publicly available tool, https://preventabledeathstracker.net/, which displays coroners’ reports in England and Wales to streamline access and identify important lessons to prevent future deaths.

**Conclusion:** National organisations, healthcare professionals, and prescribers should take actions to address the concerns of coroners’ in PFDs to improve the safe use of anticoagulants in patients with cardiovascular disease.

**How this fits in:** A previous assessment of 500 PFDs identified anticoagulants as the class of drugs most often involved in fatal medication errors. This study uses innovative methods to automatically collect all available PFDs between 2013 and 2019 to identify deaths from cardiovascular disease when the use of or lack of anticoagulants caused or contributed to the death. Coroners’ raised hundreds of coroners in their reports, including issues with communication, following protocols, education and training, access to resources, and safety. Despite repeat concerns with national relevance being identified, most CVD-anticoagulant PFDs were sent locally to NHS Trusts, hospitals, and general practices, limiting their ability to reduce harms and prevent premature deaths.

## Introduction

Cardiovascular disease (CVD) is the leading cause of mortality worldwide (1,2). In 2019, 18.6 million deaths (33% of all deaths) were due to CVD (3), with a projection of 24 million annual deaths by 2030 (4). In England and Wales, CVD was responsible for almost a quarter of all deaths in 2019 (5,6). Premature mortality from CVD in England has also been attributed to greater socioeconomic inequalities in people under 75 years of age (7).

In patients at high risk of strokes, heart attacks, deep vein thrombosis, or pulmonary embolism, anticoagulation is one possible prophylactic intervention (8). Anticoagulants target different points of the coagulation cascade, helping to prevent blood clot formation and the adverse effects of excessive clotting. In English primary care, the prescribing of anticoagulants increased from 15 million doses to 33 million between January 2014 and August 2019 (9). Three main types of anticoagulants are outlined in guidance published by The National Institute for Health and Care Excellence (NICE): low molecular weight heparin (e.g. enoxaparin), vitamin K antagonists (e.g. warfarin), and direct-acting oral anticoagulants (DOACs; e.g. rivaroxaban). The effectiveness of anticoagulants for cardiovascular disease is well established. For example, adjusted-dose warfarin reduced stroke by 62% (95% CI: 48% to 72%) in patients with atrial fibrillation (10). However, the narrow therapeutic index and frequent laboratory monitoring needed with warfarin administration have led to the development of DOACs (11). Bleeding associated with warfarin therapy is among the top three adverse drug reactions that cause hospital admissions in England (12).

Coroners’ reports, previously named Rule 43 and now called Prevention of Future Death reports (PFDs), are written when the coroner believes that action is necessary to prevent a death (13). PFDs are sent to specific individuals or organisations, who, under regulation 28 and 29 of the Coroners (Investigations) Regulations 2013, have a duty to respond within 56 days of the date of report (14). Previous analysis of coroners’ reports has shown that anticoagulants were the drugs most commonly reported to have been involved in fatal medication errors in England and Wales (15,16). This analysis also found that coroners’ most commonly raised concerns regarding adverse drug reactions to prescribed medicines, followed by omissions of necessary treatment and monitoring failures. In this study, the authors examined only a proportion (n=500) of all published coroners’ reports. Building on previous research (15,16), we aimed to assess all available PFDs between 2013 ad 2019 for deaths that involved individuals with CVD, in whom the use or lack of use of anticoagulants caused or contributed to the death. We sought to discover: 1) what concerns were highlighted by coroners; 2) to which individuals or organizations PFDs were addressed; 3) whether responses were made by the individuals or organizations to whom the PFDs were sent.

## Methods

We designed a retrospective observational study and preregistered our study protocol on an open repository (17). We used the STROBE reporting guideline to write our manuscript.

### Data collection

PFDs are openly available on the Courts and Tribunals Judiciary website (18). We used web scraping to automatically collect PFDs, and from the output created the Preventable Deaths Database and the Preventable Deaths Tracker: https://preventabledeathstracker.net/ (19). The code to create the scraper is openly available on GitHub (20) and has been previously described (21). The Preventable Deaths Database contains: the case reference number; the date of the report; the name of the deceased; the coroner’s name; the coroner’s jurisdiction; the category of death (defined by the Chief Coroners’ office); to whom the report was sent; and the URL to the Judiciary website. For population data on deaths from CVD, we used the most recent (2001-2019) dataset of deaths registered in England and Wales, released by the Office for National Statistics in 2020 (ONS) (5).

### Eligibility of cases

We examined all cases (n=3037) in the Preventable Deaths Database from July 2013 (the first date on which they were uploaded to the Judiciary website) to December 2019. We screened the cases independently in duplicate (AA & GCR) to determine whether CVD caused or contributed to the death using a pre-defined algorithm (Supplementary Figure 1). We used Chapter IX (Diseases of the Circulatory System) of the International Statistical Classification of Diseases and Related Health Problems 10th Revision (ICD-10) to align with the ONS classifications of death. When the deceased suffered from a single condition listed under chapter IX that could not be unequivocally attributed to external causes, the case was included. We then screened for cases where one or more anticoagulant caused or contributed to the death or where the coroner suggested that had an anticoagulant been given, it would have prevented the death. We defined anticoagulants as agents targeting different points of the coagulation pathway to prevent clot formation. Discrepancies were resolved by consensus discussion (AA, GCR, & JKA).

### Data extraction

For included cases, one study author (AA) manually extracted the following variables into a predesigned Google Sheet, which was cross examined by another study author (GCR): the individuals or organizations to whom reports were sent, who responded, the due date of response and the date received; date of death; the dates on which the inquest started and ended; age; sex; setting or location of death; medical cause(s) of death; the coroner’s conclusion(s) of the inquest; relevant medical, mental health, and social history; whether any substance(s) were implicated in the death and the type of substance(s); coroner’s concern(s) and actions proposed by the coroner.

**Figure 1:**
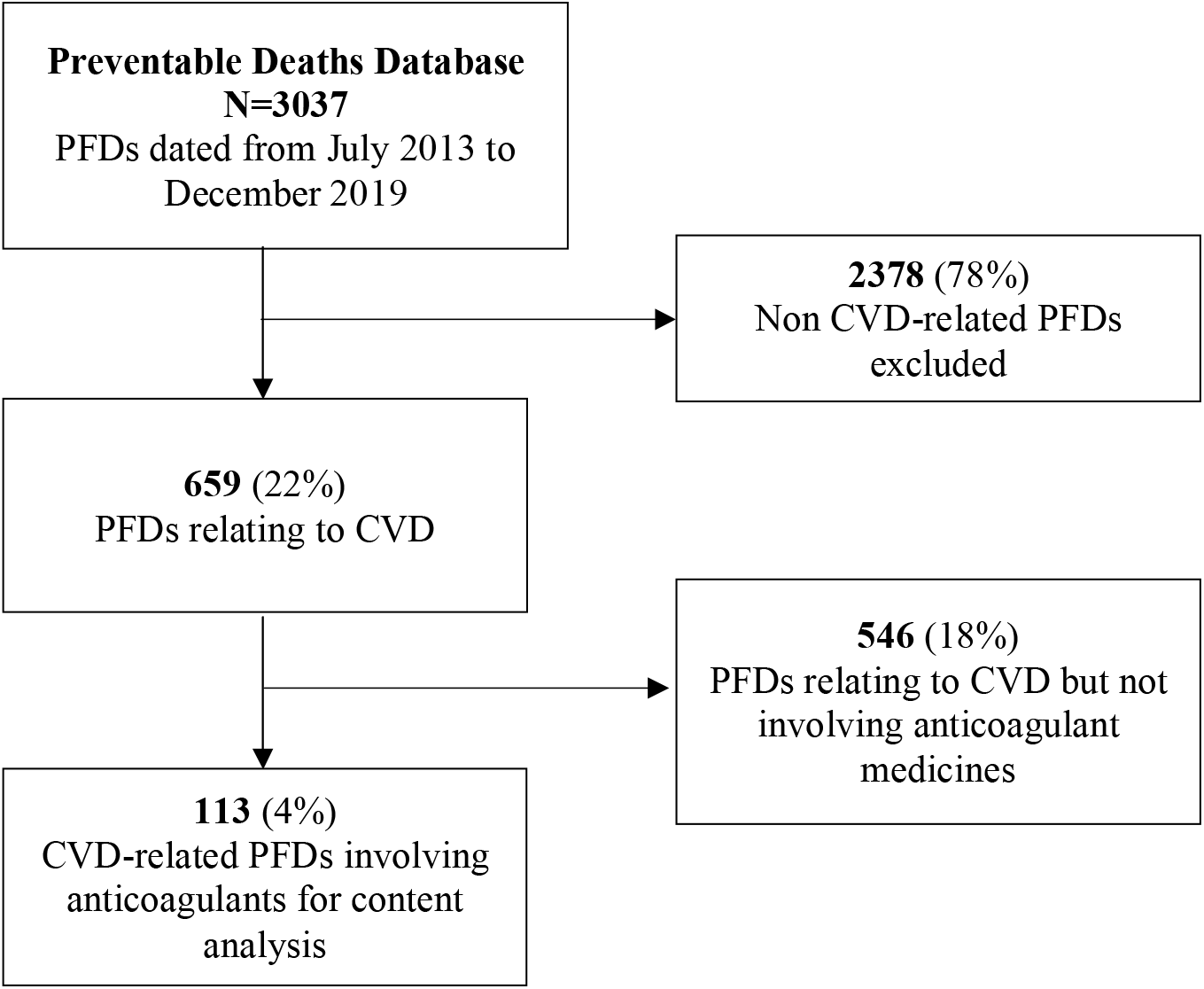
Flow diagram showing the numbers of Prevent Future Death reports (PFDs) included and excluded from the Preventable Deaths Database using the eligibility criteria for this study

### Data analysis

To determine the annual deaths from CVD in England and Wales, we filtered the ONS data (5) for deaths caused by conditions listed under chapter IX of ICD-10 (ICD-10 codes: I00-I99). We summed the number of reports written each year and compared the totals with ONS mortality data for CVD. We used descriptive statistics to report the quantitative findings and performed content analysis (22) to classify the concerns and actions raised by coroners with categories derived inductively. We calculated a response rate for each organisation as the proportion of reports to which a response was submitted over the total number received. Responses were further classified as either on-time (delivered within 56 days), late (submitted after 56 days), or overdue (when no response was found on the Judiciary website). A response rate of 100% means that individual or organisation adhered to regulation 28 of the Coroners’ (Investigations) Regulations 2013 and responded to all PFDs issued by coroners.

### Software and data sharing

We used Python v3.7 in Jupyter Notebooks with pandas, seaborn, and matplotlib libraries to analyse the data and create figures. The data, statistical code, and study materials are openly available via the Open Science Frame (23) and GitHub (24).

## Results

In 659 cases (22% of all PFDs) CVD caused or contributed to the death (Figure 1). Of the CVD-related PFDs, 17% (n=113) involved or mentioned the use or lack of an anticoagulant. Over the seven-year study period, there was a median of 16 (IQR: 15-17) CVD-related anticoagulant PFDs each year (Supplementary Table 1).

In 99 cases, an anticoagulant caused or contributed to the death. Warfarin (36%) was the most common anticoagulant specified, followed by enoxaparin (11%) and rivaroxaban (11%) (Figure 2). In 14 cases, the coroner mentioned that the administration of an anticoagulant might have prevented the death (Figure 2). There were equal proportions of males (n=56) and females (n=57) in the 113 cases. The median age of the deceased were 76 years (IQR: 61-84 years; n=77).

**Figure 2:**
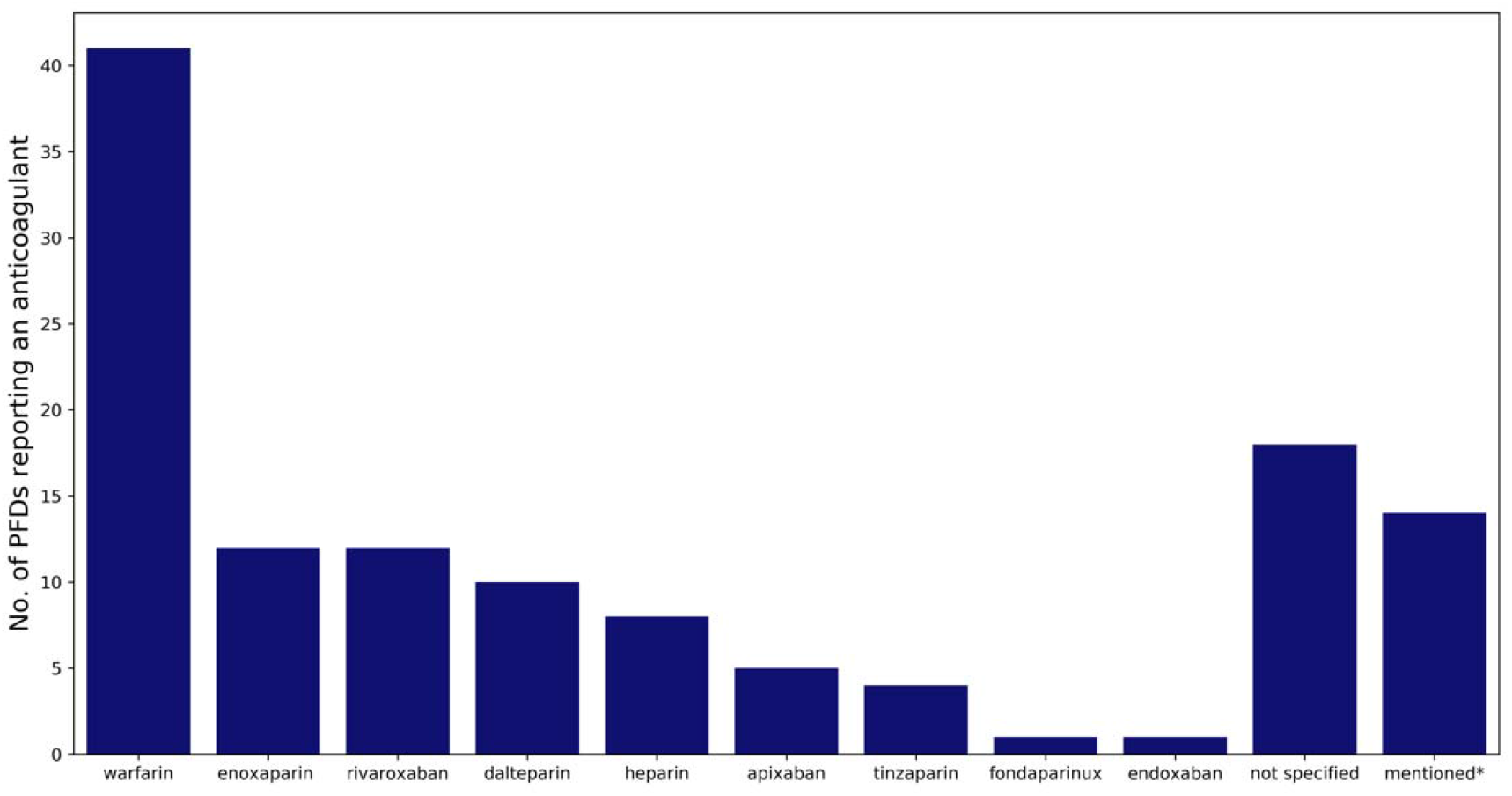
Anticoagulants reported in cardiovascular disease-related Prevent Future Death reports (PFDs). *in these PFDs, the coroner suggested that if an anticoagulant had been used the death might have been prevented

Seventy-five coroners across 36 jurisdictions wrote 113 PFDs. Coroners in the North West (25%) and South East (19%) of England wrote the most, whereas those in the East (2%) and North East (3%) of England wrote very few (Table 1).

**Table 1:**
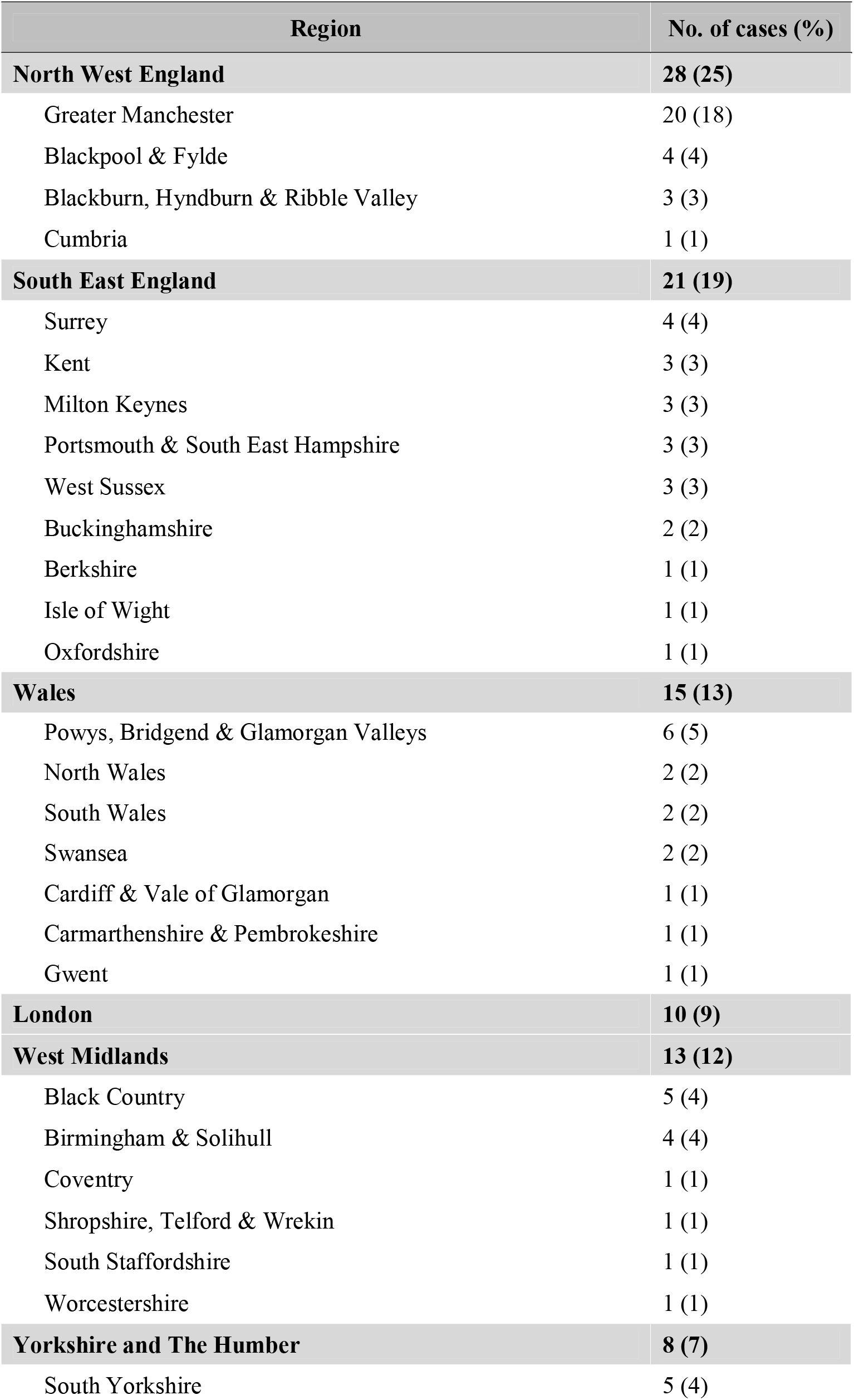
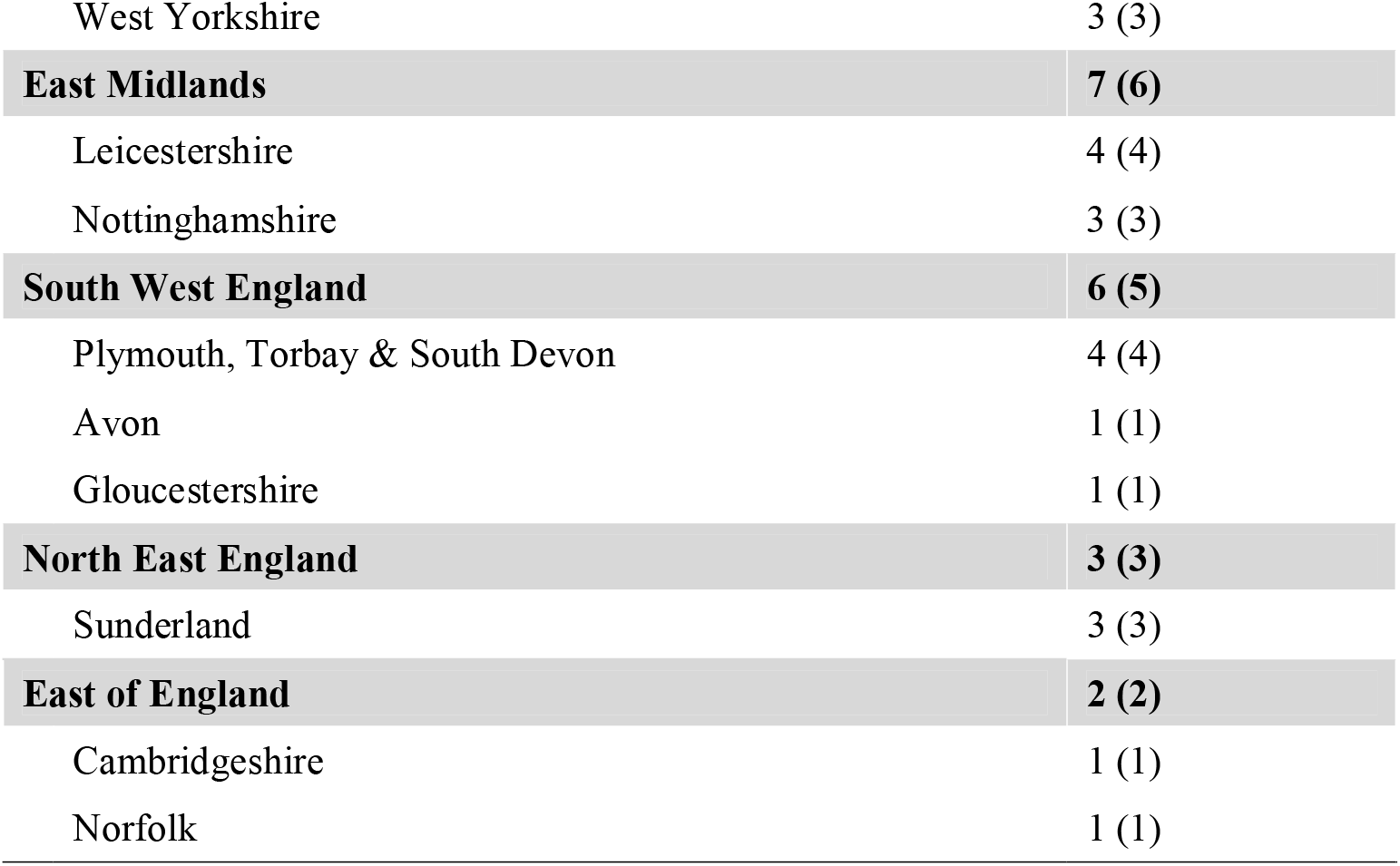
Jurisdictions of coroners who issued the 113 Prevent Future Death reports involving an anticoagulant and cardiovascular disease

We identified 335 individual concerns raised by coroners in the 113 cases. Using content analysis, we categorised these concerns into 51 groups and five higher-order themes, including communication, failure to follow protocols, education and training, resources, and safety (Table 2). The most common concerns were poor systems (31%), poor communication (25%), failure to keep accurate medical records (25%), and failures or delays in having appropriate assessments done (17%). Concerns most frequently belonged to the theme of following protocols (36%), followed by communication (22%), and safety issues (21%).

**Table 2:**
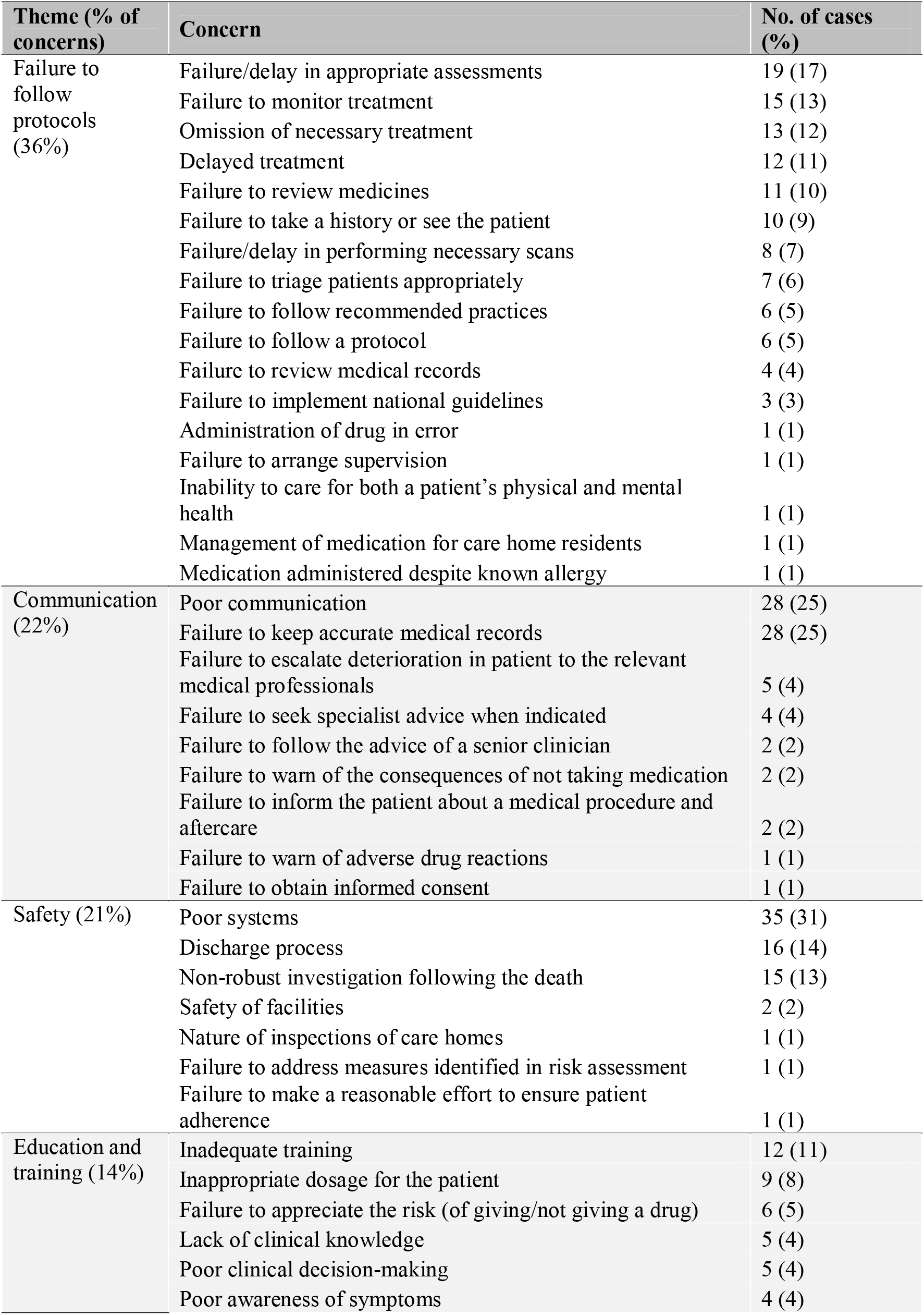
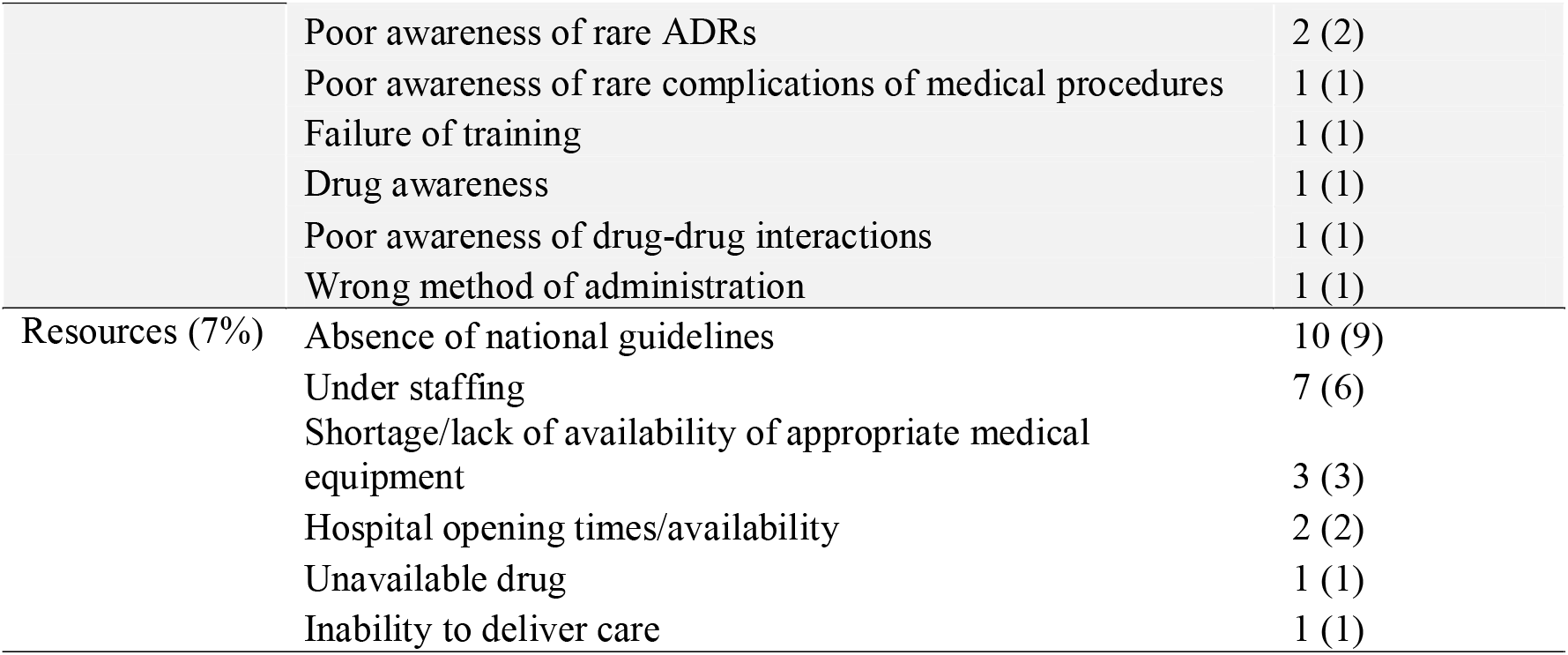
Concerns raised by coroners, grouped by five higher-order themes, and how often they were reported

In 82% of CVD-related PFDs involving anticoagulants, coroners stated that “action should be taken” (Supplementary Table 2). When coroners suggested further actions, we grouped their actions into 28 categories. Ensuring effective communication (3%), introducing new policies and protocols (2%), and reviewing the handling of prescriptions (2%) were the most common actions proposed (Supplementary Table 2).

For the 113 CVD-related PFDs involving anticoagulants, coroners sent 181 reports to 37 individuals and organisations (Table 3). Local services such as NHS Trusts (29%), hospitals (10%), and general practices (8%), were addressed PFDs most frequently. By statute addressees must respond to the coroner within 56-days, but only 29% responded on time; 11% responded late and 60% were overdue. Medical societies (0%) and royal colleges (0%) had the lowest response rates, while NHS entities had the highest, albeit with half of their responses overdue. Ranking recipients by response rate and response time, NHS 111, NHS Wales, and CCGs performed best (Supplementary Table 3).

**Table 3:** Recipients of coroners Prevent Future Death reports (PFDs) and their response rates to reports

| Addressee | No. of reports received | No. of responses | Response rate (%) | Classification of responses |  |  |
| --- | --- | --- | --- | --- | --- | --- |
|  |  |  |  | % On time | % Late | % Overdue |
| **NHS entities** | **118** | **60** | **51** | **38** | **13** | **49** |
| CCGs | 3 | 3 | 100 | 67 | 33 | 0 |
| NHS 111 | 1 | 1 | 100 | 100 | 0 | 0 |
| NHS Wales | 1 | 1 | 100 | 100 | 0 | 0 |
| Ambulance services | 7 | 5 | 71 | 57 | 14 | 29 |
| NHS Trusts | 53 | 32 | 60 | 45 | 15 | 40 |
| NHS England | 4 | 2 | 50 | 50 | 0 | 50 |
| hospitals | 19 | 7 | 37 | 26 | 11 | 63 |
| General practices | 14 | 5 | 36 | 29 | 7 | 64 |
| Local Health Boards | 3 | 1 | 33 | 0 | 33 | 67 |
| University Health Boards | 10 | 3 | 30 | 20 | 10 | 70 |
| Mental Health Trusts | 2 | 0 | 0 | 0 | 0 | 100 |
| NHS Pathways | 1 | 0 | 0 | 0 | 0 | 100 |
| **Government** | **17** | **5** | **29** | **18** | **12** | **71** |
| DHSC | 7 | 3 | 43 | 14 | 29 | 57 |
| Local authorities | 3 | 1 | 33 | 33 | 0 | 67 |
| Welsh Government | 7 | 1 | 14 | 14 | 0 | 86 |
| **Professional bodies** | **21** | **4** | **19** | **10** | **10** | **81** |
| CQC | 6 | 2 | 33 | 17 | 17 | 67 |
| NICE | 8 | 2 | 25 | 13 | 13 | 75 |
| GMC | 2 | 0 | 0 | 0 | 0 | 100 |
| BMA | 1 | 0 | 0 | 0 | 0 | 100 |
| GDC | 1 | 0 | 0 | 0 | 0 | 100 |
| MHRA | 1 | 0 | 0 | 0 | 0 | 100 |
| AACE | 1 | 0 | 0 | 0 | 0 | 100 |
| The Renal Association | 1 | 0 | 0 | 0 | 0 | 100 |
| **Others** | **18** | **3** | **17** | **11** | **6** | **83** |
| Police | 1 | 1 | 100 | 0 | 100 | 0 |
| Private companies | 4 | 1 | 25 | 25 | 0 | 75 |
| Care homes | 9 | 1 | 11 | 11 | 0 | 89 |
| Carewatch | 1 | 0 | 0 | 0 | 0 | 100 |
| Highway maintenance | 1 | 0 | 0 | 0 | 0 | 100 |
| Housing Associations | 1 | 0 | 0 | 0 | 0 | 100 |
| Local Charities | 1 | 0 | 0 | 0 | 0 | 100 |
| **Medical societies** | **4** | **0** | **0** | **0** | **0** | **100** |
| BCS | 1 | 0 | 0 | 0 | 0 | 100 |
| BRS | 1 | 0 | 0 | 0 | 0 | 100 |
| ICS | 1 | 0 | 0 | 0 | 0 | 100 |
| RPS | 1 | 0 | 0 | 0 | 0 | 100 |
| **Medical royal colleges** | **3** | **0** | **0** | **0** | **0** | **100** |
| RCGP | 1 | 0 | 0 | 0 | 0 | 100 |
| RCOG | 1 | 0 | 0 | 0 | 0 | 100 |
| RCP | 1 | 0 | 0 | 0 | 0 | 100 |
| **Total** | **181** | **72** | **40** | **29** | **11** | **60** |

AACE: Association of Ambulance Chief Executives; BCS: British Cardiovascular Society; BMA: British Medical Association; BRS: British Renal Society; CQC: Care Quality Commission; DHSC: Department of Health and Social Care; GDC: General Dental Council; GMC: General Medical Council; ICS: Intensive Care Society; MHRA: Medicines and Healthcare products Regulatory Agency; NICE: The National Institute for Health and Care Excellence; NHS: National Health Service; RCGP: Royal College of General Practitioners; RCOG: Royal College of Obstetricians and Gynaecologists; RCP: Royal College of Physicians; RPS: Royal Pharmaceutical Society

## Discussion

### Summary

We identified 113 premature deaths from cardiovascular disease involving anticoagulants. In 88% of cases, the use of one or more anticoagulant resulted in death. In 12% of cases, the administration of an anticoagulant may have prevented death. We found wide geographical variation in the issuing of PFDs and the type of information reported, with coroners in Greater Manchester writing the most. Coroners raised hundreds of concerns, some relating directly to the risks of anticoagulation and the caution with which patients on these drugs should be managed, though rarely were the concerns not already mentioned in guidance available to healthcare professionals. Most of these concerns were addressed locally and under regulation 28, 109 individuals or organisations were overdue in their response to coroners.

### Strengths and limitations

We used a reproducible data-collection method to examine all available PFDs from inception to 2019, which provided an estimated 40-fold time saving (21), and reduced the potential for selection bias. However, the PFDs included in our study depend on the working practices of coroners and the Chief Coroner’s Office in uploading PFDs and their responses to the website. The 113 PFDs cannot therefore represent all preventable deaths from CVD involving anticoagulants in England and Wales. There were also missing data; for example, 32% of PFDs did not report age and 16% did not specify the type of anticoagulant; this may be attributed to lack of PFD training provided to coroners. Our findings are also limited by the available data and information provided by coroners in PFDs, thus it is not possible to examine the relationship between cardiovascular disease and anticoagulants in causing death, or to differeintiate between the appropriate use, misuse, and underuse of anticoagultants.

### Comparison with existing literature

Our study builds on prior research that evaluated smaller samples of coroners’ reports (15,16,25–27). Compared with previous studies (26,28), we did not identify a sex imbalance. However, we found that new hazards were rarely identified and that most PFDs were addressed locally, as shown by Ferner et al. (15), which means that valuable lessons were not widely disseminated and may be why we found that coroners repeatedly expressed similar concerns, in line with former research (25). This questions whether PFDs are fulfilling their purpose. Similar to Fox and Jacobson (26), we found pronounced geographical variation in the issuing of PFDs and poor response rates. A review of the coronial system in England and Wales highlighted a lack of accountability, leadership, and quality assurance (29). Since there is no system in place for enforcing or auditing compliance with regulation 28 or assessing the quality of PFDs and the adequacy of responses and actions taken to prevent deaths, our findings show that the system has scope for improvement.

General practices were sent the third highest number of CVD-related PFDs involving anticoagulants, but collectively only 36% responded. This may be because of a lack of awareness of the statutory requirements and medico-legal training of GPs, as identified by previous research (30). During the COVID-19 pandemic, GPs called for deaths of colleagues to be reported to the coroner and PFDs to be issued (31). Fortunately, the value of PFDs as a tool for improving clinical practice is being recognised, and efforts are underway to widely disseminate their lessons to healthcare professionals, policymakers, and the public (32–37).

### Implications for research and/or practice

Concerns raised by coroners provide lessons for prescribers and policymakers on the safety and proper use of therapies. During the COVID-19 pandemic, patients were switched from warfarin to other oral anticoagulants, given the need for less frequent blood testing (38). Drugs such as andexanet alfa could prove critical in the outcome of patients with severe bleeding during treatment with apixaban or rivaroxaban. Since the coroner expressed concerns (Case: 2018-0032), NICE has drafted an appraisal for andexanet alfa (39), and its publication is expected on 12 May 2021 (40). There has also been a phased launch of andexanet alfa in UK hospitals (41). However, it is unclear whether this was a direct result of the PFD, since no response from the MHRA was published on the Judiciary website.

When PFDs are addressed to the appropriate recipients at the national level, their actions can help prevent deaths. For example, a PFD was sent to NICE when the deceased suffered a fall while taking an anticoagulant without having the appropriate neuroimaging performed (Case: 2019-0106). The coroner’s concerns were acknowledged in a NICE surveillance report (42) and resulted in updating of the NICE guideline on head injury, to emphasise that people taking DOACs should be investigated with the same care as those taking warfarin (43). However, we also identified unaddressed concerns about national guideline; three-quarters of PFDs sent to NICE have no responses listed on the Judiciary website. Restarting warfarin after a head injury is particularly important, as delays could leave the patient at risk of a stroke, but resuming too soon may lead to haemorrhage. A retrospective review of the medical charts of 256 patients admitted to a trauma centre in West Texas between 2009 and 2012 showed that patients who resumed anticoagulant therapy at 7-10 days after the injury had the best prognosis (44). PFDs can therefore be used to update guidance and inform future prospective cohort studies.

Our study provides a database and resource (https://preventabledeathstracker.net/) for future evaluations of PFDs. Further content analysis should be used to assess the 181 responses to the 113 PFDs, to assess the adequacy of actions proposed to prevent deaths and their implementation. Future research could use our open data to examine coroners’ concerns in the 546 CVD-related PFDs not involving anticoagulants and their responses. In building the web scraper to collect PFDs, we found various inconsistencies and omissions on the Judiciary website, which should be addressed to improve the quality of data. The missing data from PFDs we have highlighted reveal target areas for coronial training in the writing of PFDs.

### Conclusions

This study used sophisticated, reproducible, and internationally recognised data-collection methods (21) to demonstrate that PFDs provide valuable lessons when prescribing anticoagulants and managing patients with CVD. However, it is unclear whether actions are being taken to incorporate such lessons. To improve patient safety, lessons should be widely disseminated and used in practice.

## Data Availability

Study materials, protocol, data, and statistical code are openly available on the OSF (https://osf.io/qkfjp/) and GitHub (https://github.com/georgiarichards/CVD_anticoags)

## Ethical approval

As the case reports are publicly available on the Judiciary website, approval from an ethics committee was not required.

## Acknowledgements and contributions

AA proposed the topic for research, contributed to the protocol and devised the algorithm for the eligibility of cases, screened the database for eligible case reports, extracted and analysed the data, and wrote the first draft of the manuscript. GCR designed the study, wrote the protocol, sourced the data, contributed to the web scraper, provided data management, dual screened the database for eligible cases, analysed the data, provided primary supervisory support and substantiality contributed to subsequent manuscript drafts. JKA and CH reviewed the protocol and preliminary findings and provided supervisory support. NJD wrote the code and advised on the web scraper. All authors have full access to all the data in the study, read and approved the final manuscript, and accept responsibility to submit for publication.

