## Supplementary material for "Deaths from cardiovascular disease involving anticoagulants: a systematic synthesis of coroners’ case reports": Figure S1 in Supplement

### Supplementary materials

**Figure S1:** Algorithm for the selection of cardiovascular disease-related PFDs involving anticoagulants

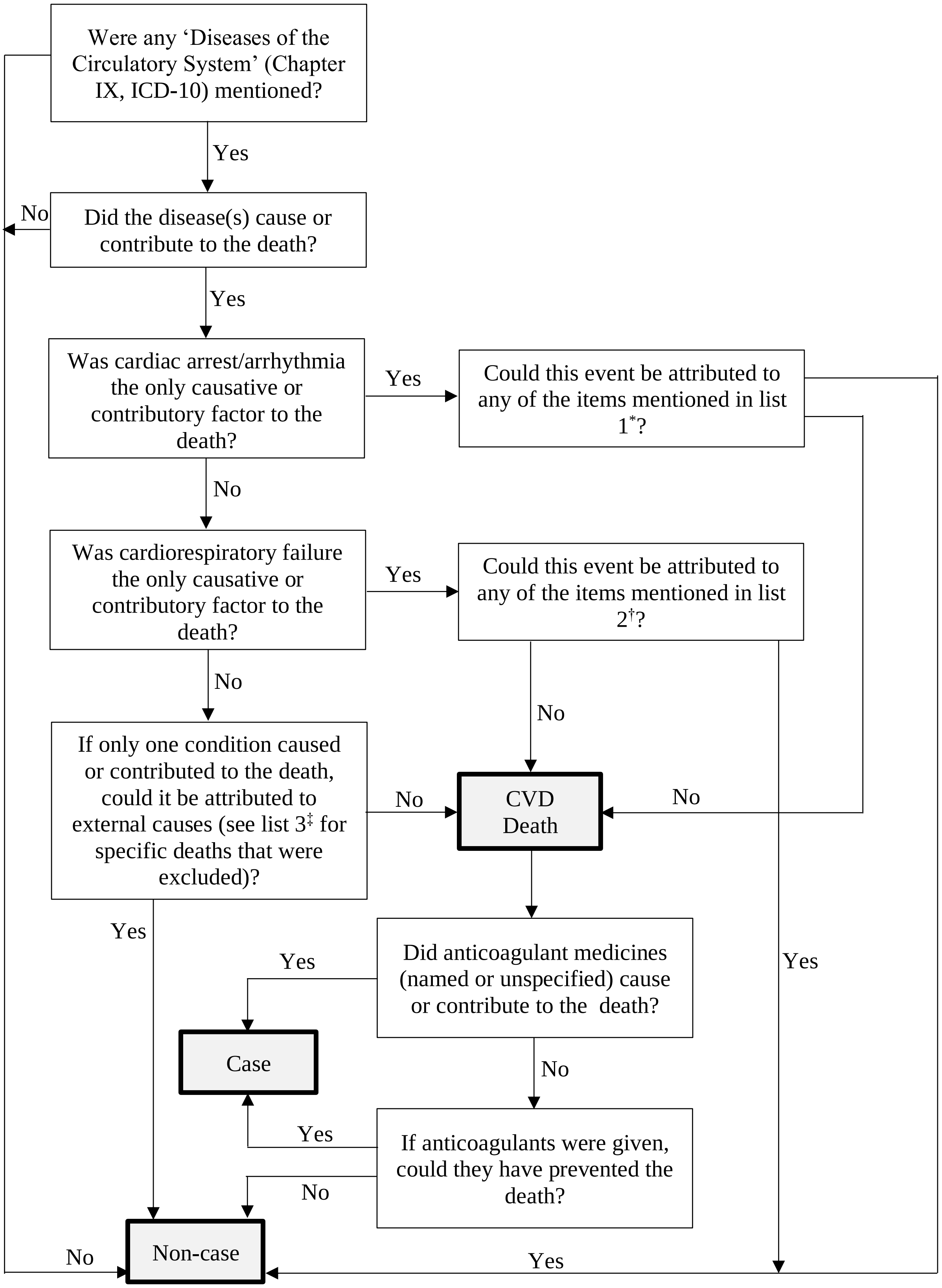

[*]List 1: prolonged hypoxia; hypoxia caused by impaired respiratory function; occlusion of the airways; positional asphyxia; covering applied over the head; oxygen supply not being connected properly; failed attempt at intubation; choking; failure to carry out prompt ventilation after aspiration; a synthetic cannabinoid; reaction to a drug; presence of drugs (according to the pathologist's ruling); clozapine associated myocarditis; general anaesthetic therapies; ingestion of E-cigarette fluid; exposure to a known allergy; car collision; a fall (traumatic cardiac arrest); persistent epistaxis; suicide/ hanging; seizure; sepsis; legionella pneumonia; Addisonian crisis; electrolyte disturbance (e.g. hyperkalaemia); intussception of the bowel; vomiting caused by an intermittent blockage of the bowel; ectopic pregnancy; congenital malformations of the great artery

[†]List 2: prone position; bronchopneumonia; fire; ileus of the small intestine, due to restoration of bowel continuity which in turn was due to Crohn's disease; severe anaemia due to sickle cell disease

[‡]List 3: haemorrhage resulting from a fall; intraabdominal haemorrhage following medical intervention (colonscopy); injuries to blood vessels resulting from bad surgical planning; injury of heart and pericardium; gastrointestinal bleeding due to deep duodenal ulcer; congestive cardiac failure resulting from inadvertent fluid overload of Total Parenteral Nutrition (TPN) given via an UVC; cardiac tamponade as complication of central lines being put in for parental nutrition; cerebral infarct following cannulation of the carotid artery rather than the vein; medical procedure penetrated the patient's heart causing it to stop; medical procedure caused acute arteritis of innominate artery; air embolization and cerebral infarction resulting from the clamp on a central venous catheter being left open; laparoscopy which caused a leaking oesophagus leading to mediastinitis, compromised breathing and cardiac arrest; stroke as a complication of a liver transplant; pseudoaneurysm of the left thigh which developed as a result of intravenous drug abuse; multiple complications of a cardiac trial

**Table S1:** Cardiovascular disease-related Prevent Future Death reports in comparison with the Office of National Statistics mortality data and those involving anticoagulants by year

| Year | Total no. of PFDs | No. of CVD-related PFDs | No. of CVD-related PFDs involving anticoagulants | No. of CVD deaths reported to the ONS | % of CVD-related PFDs vs ONS CVD deaths |
|---|---|---|---|---|---|
| 2013 | 173 | 34 | 7 | 140301 | 0.024 |
| 2014 | 555 | 133 | 17 | 135904 | 0.098 |
| 2015 | 477 | 105 | 26 | 138614 | 0.076 |
| 2016 | 470 | 99 | 14 | 133705 | 0.074 |
| 2017 | 435 | 108 | 16 | 133511 | 0.081 |
| 2018 | 413 | 81 | 16 | 132233 | 0.061 |
| 2019 | 514 | 99 | 17 | 129421 | 0.076 |
| **Total** | 3037 | 659 | 113 | 943689 | 0.070 |
| **Median (IQR)** | 470 (424-495) | 99 (90-107) | 16 (15-17) | 133705 (132872-137259) | 0.075 (0.068-0.078) |

CVD: cardiovascular disease; IQR: interquartile range; ONS: Office of National Statistics; PFDs: Prevention of Future Deaths reports

**Table S2:** Actions recommended by coroners and the number of reports in which they featured

| Recommendations | No. of cases (%) |
|---|---|
| Action should be taken | 93 (82) |
| Ensure effective communication | 3 (3) |
| Introduce new policy and protocols | 2 (2) |
| Review handling of prescriptions | 2 (2) |
| Review medical record-keeping | 2 (2) |
| Review national guidelines | 2 (2) |
| Review policy and protocols | 2 (2) |
| Review procedures for risk assessments | 2 (2) |
| Review training of relevant staff | 2 (2) |
| Carry out further inspections | 1 (1) |
| Ensure accurate record keeping | 1 (1) |
| Ensure medical records are available electronically | 1 (1) |
| Ensure patients are reviewed promptly | 1 (1) |
| Ensure process exists to report patient non-compliance | 1 (1) |
| Ensure recommended practice is followed | 1 (1) |
| Ensure sufficient staffing levels | 1 (1) |
| Improve booking system | 1 (1) |
| Improve communication | 1 (1) |
| Improve medical record-keeping | 1 (1) |
| Provide training | 1 (1) |
| Review alarm system | 1 (1) |
| Review discharge process | 1 (1) |
| Review fitness to practice | 1 (1) |
| Review IT systems | 1 (1) |
| Review methods of communication | 1 (1) |
| Review referral process | 1 (1) |
| Review service provision | 1 (1) |
| Review treatment of current patients | 1 (1) |

**Table S3:** Addressees' responses to coroners' Prevention of Future Death reports based on response rate and responding on-time, within 56 days from the date of the report

| Rank | Addressee | Response rate (%) | % On-time | % Late | % Overdue |
|---|---|---|---|---|---|
| 1 | NHS 111 | 100 | 100 | 0 | 0 |
| 1 | NHS Wales | 100 | 100 | 0 | 0 |
| 2 | CCGs | 100 | 67 | 33 | 0 |
| 3 | Police | 100 | 0 | 100 | 0 |
| 4 | Ambulance | 71 | 57 | 14 | 29 |
| 5 | NHS Trusts | 60 | 45 | 15 | 40 |
| 6 | NHS England | 50 | 50 | 0 | 50 |
| 7 | DHSC | 43 | 14 | 29 | 57 |
| 8 | Hospitals | 37 | 26 | 11 | 63 |
| 9 | General practices | 36 | 29 | 7 | 64 |
| 10 | Local authorities | 33 | 33 | 0 | 67 |
| 11 | CQC | 33 | 17 | 17 | 67 |
| 12 | Local Health Board | 33 | 0 | 33 | 67 |
| 13 | University Health Board | 30 | 20 | 10 | 70 |
| 14 | Private company | 25 | 25 | 0 | 75 |
| 15 | NICE | 25 | 13 | 13 | 75 |
| 16 | Welsh Government | 14 | 14 | 0 | 86 |
| 17 | Care home | 11 | 11 | 0 | 89 |
| 18 | AACE | 0 | 0 | 0 | 100 |
| 18 | BCS | 0 | 0 | 0 | 100 |
| 18 | BMA | 0 | 0 | 0 | 100 |
| 18 | BRS | 0 | 0 | 0 | 100 |
| 18 | Carewatch | 0 | 0 | 0 | 100 |
| 18 | GDC | 0 | 0 | 0 | 100 |
| 18 | GMC | 0 | 0 | 0 | 100 |
| 18 | Highway maintenance | 0 | 0 | 0 | 100 |
| 18 | Housing Association | 0 | 0 | 0 | 100 |
| 18 | ICS | 0 | 0 | 0 | 100 |
| 18 | Local Charity | 0 | 0 | 0 | 100 |
| 18 | Mental Health trust | 0 | 0 | 0 | 100 |
| 18 | MHRA | 0 | 0 | 0 | 100 |
| 18 | NHS Pathways | 0 | 0 | 0 | 100 |
| 18 | RCGP | 0 | 0 | 0 | 100 |
| 18 | RCOG | 0 | 0 | 0 | 100 |
| 18 | RCP | 0 | 0 | 0 | 100 |
| 18 | RPS | 0 | 0 | 0 | 100 |
| 18 | The Renal Association | 0 | 0 | 0 | 100 |

AACE: Association of Ambulance Chief Executives; BCS: British Cardiovascular Society; BMA: British Medical Association; BRS: British Renal Society; CQC: Care Quality Commission; DHSC: Department of Health and Social Care; GDC: General Dental Council; GMC: General Medical Council; ICS: Intensive Care Society; MHRA: Medicines and Healthcare products Regulatory Agency; NICE: The National Institute for Health and Care Excellence; NHS: National Health Service; RCGP: Royal College of General Practitioners; RCOG: Royal College of Obstetricians and Gynaecologists; RCP: Royal College of Physicians; RPS: Royal Pharmaceutical Society
